# The Prognostic Value of Genetic Architectures in Cognitive Decline

**DOI:** 10.64898/2026.07.13.26357971

**Authors:** Michael J. Espero

## Abstract

**Background & Methods:** The multifaceted physical nature of heritable cognitive impairment in dementia presents significant challenges for traditional linear frameworks attempting to model synergistic risk. While various loci are identified as contributing to neurocognitive disparities, the emergent phenotypic expression and associated predictive value relative to standard clinical baselines require further investigation. To facilitate dimensional reduction of complex genetic data into identifiable phenotypes, Generalized Low Rank Modeling (GLRM) and K-means clustering are applied to participant data from the Alzheimer’s Disease Neuroimaging Initiative (ADNI). The utility of these derived archetypes and clusters is assessed, stratifying variance for Mini-Mental State Examination (MMSE) performance. Utilizing generalized additive modeling (GAM) and partial eta squared (*η_p_*^2^) effect size, the derived genetic features are compared with other predictors including age, educational attainment, gender, and raw, genetic variant carriage dimensions.

**Results & Conclusion:** In accordance with the hypothesized empirical regularity, age and education persist as primary predictors of MMSE performance. The unsupervised machine learning pipeline successfully identified a composite genetic cluster that emerged as an influential predictor in terms of relative magnitude (*η_p_*^2^). Centroid analysis of the GLRM subspace indicated that a particular sub-population (Cluster 2) - defined by a substantial weighting on the *EPHA1* target - demonstrated a statistically significant association with MMSE scores, relative to cluster 3. These results suggest that data-driven genetic feature engineering provides an interpretable basis for inference regarding variance in global cognition. By discovering multivariate genetic architecture, this modeling approach captures complexity *often missed* by individual clinical variable modeling. Such findings implicate the utility of interpretable machine learning for translational dementia research and predictive clinical stratification.

## Introduction

A characterizing feature of dementias such as Alzheimer’s (AD) is often distinguished as deficit in episodic memory (Bangert & Balota, 2012; Ferencz, Laukka, Welmer, Kalpousos, Angleman, Keller, Graff, Lovden, and Backman 2014; Greenbaum et al., 2016). Impairment in verbal memory, executive function, and other complex cognitive abilities appear to coincide (Salthouse & Becker, 1998). Heritable variance in facets of neurocognition may influence specific disparities across the human lifespan. The notion that between-person differences in genes can account for distinctive aspects of cognitive abilities implicates the importance of the physical nature from which mental faculties emerge. The carriage of specific genetic variants may support, impede, or be indifferent (as sampled and measured) to known neurocognitive pathways (i.e. superior longitudinal fasciculi) and influence functional domains widely associated with certain dementias. In this sense, certain genetic variants may predispose individuals to cognitive impairment, dementia, and associated risk for downstream accumulation of plaques, tangles, disruption of neural networks, and genuine deficits in cognitive performance.

### Single Nucleotide Polymorphisms (SNPs)

**Clusterin (CLU)** proteins exhibit context-specific roles in prevention of misfolding (“chaperone” function), the transport of blood fats, trafficking intercellular aggregations, as well as regulation of cell survival and death (Foster, 2019). CLU variants suggest differential effects in terms of neurocognitive disease risk and cognitive performance. Cai et al. (2016) found carriers of the CLU (rs11136000) TT genotype at reduced risk for mild cognitive impairment (MCI) relative to CC carriers after adjustment for age, educational attainment, and gender, OR = 0.158, χ^2^ = 4.113, *p* = 0.043. CLU’s (rs11136000) influence on fluid reasoning (Gf), processing speed (Gs), verbal fluency, and working memory (WM) performance may be pronounced, as TT carriers performed better (TT genotype as cognitively beneficial) than CC and heterozygotes (CT) with regard to trail-making task (TMT) speededness, a cognitive assessment thought to tap Gf and Gs (Salthouse, 2011), TT: 168.6 (± 91), CC and CT: 244.6 (± 112.7), *p* = 0.023. TT carriers performed better in assessment of verbal fluency, TT: 11 (± 5.1), CC and CT: 7.5 (± 4.3), *p* = .012. Moreover, TT carriers performed better on the backwards digit span (EF and WM), TT: 5.3 (± 1.8), CC and CT: 3.9 (± 1.3), *p* = .021. Cruz-Sanabria (2021) observed relatively poorer performance on a third of a battery of memory tasks in CC carriers, including Reverse Verbal Span and the Free and Cued Selective Reminding Test (FCSRT), an assessment considered appropriate for detection of early stage AD and differentiation from other dementias due to its effectiveness in distinguishing *apparent* memory deficits (those associated with normal aging) from genuine memory impairments (Grober et al., 1988). Interestingly, even the benefits of the widely-promoted Mediterranean diet may differ by CLU allelic carriage. Martinez-Lapiscina’s (2014) diet experiments showed improved Mini-Mental State Exam (MMSE) scores relative to controls for carriers of the CLU (rs11136000) T allele, but not for those of other allelic carriage, *B* = 0.97, 95% CI [0.45, 1.49], *p* < 0.001. This effect remained statistically significant after adjustment for multiple comparisons.

**Phosphatidylinositol-binding clathrin assembly protein (PICALM)** plays a role in the trafficking of intracellular substrates such as lipids, neurotransmitters, and proteins (Sun, 2017). While several PICALM variants may be associated with the development of AD, the single-nucleotide polymorphism rs3851179 appears to have the strongest relationship with AD as demonstrated by replications across various ethnic cohorts including Caucasians and Chinese groups. Sun et al. (2017) showed differences by genotype in terms of brain network (default-mode network) functional connectivity, but not in performance during cognitive tasks as assessed. Barabash et al. (2013) found variants of PICALM (rs3851179) to vary with semantic fluency performance as GG carriers performed worse than AA and AG carriers, GG: 7.6 (± 4.6), AA and AG: 9.3 (± 4), *p* = .027.

**BIN1** was previously identified as a risk factor in the development of AD due to its role in modulation of tau pathologies (Dourlen, 2017). Franzmeier et al. (2019) showed increased global AV1451 tau-PET uptake among carriers of the G risk allele relative to participants with other alleles while controlling for education, gender, age, ApoE ε4 carriage, diagnosis, and gray matter (GM) volume of the global tau region of interest, *F*(81, 7) = 7.69, *d* = 0.56, *p* = 0.007. While a negative association between BIN1 rs744373 C and memory composite scores (ADNI-MEM) were previously shown, β = −0.25, *p* = 0.03, this effect was not statistically different from zero when global AV1451 tau-PET uptake was included as a mediator, β = −0.17, *p* = 0.15. Vivot (2015) found change in global cognition as measured by the MMSE negatively associated with BIN1 (rs744373) G among French participants (N = 9,294) over the age of 65, β = − 0.10, 95% CI [−0.19, −0.01]. In another sample, CC carriers performed 25% lower on average with regard to the Boston naming task relative to heterozygotes and homozygotic T carriers (Cruz-Sanabria et al., 2021). Despite increased global hippocampal volume (perhaps indicative of compensatory development), GG carriage was associated with relatively lower functional connectivity between the right DLPFC and hippocampus as well as lower working memory as assessed with increasing difficulty in later dual n-back trials (Zhang, 2015).

**Ephrin type-A receptor 1 (EPHA1)** is part of the ephrin receptor branch of the tyrosine kinase family (Wang et al., 2015) and plays a role in function of the immune system, neurotransmission, as well as other critical processes involved in the maintenance of cell membranes. Wang et al. (2015) observed statistically significant volumetric differences by genotype of brain regions thought to contribute to canonical developments in AD. At baseline, AA carriage in subjects with AD was shown to be associated with increased volume of both the right lateral occipitotemporal gyrus and right inferior temporal gyrus relative to GG and AG carriage, respectively, *p* < 0.001, *p* = 0.042.

**Sortilin receptor 1 (SORL1)** appears to influence the production of amyloid beta in its role as a precursor to proteins involved in sorting colocalization at compartmental membranes (Bralten, 2011). SORL1 carriers of several key SNPS were shown to negatively associate with global hippocampal volume. Cruz-Sanabria (2021) found improved planning and problem-solving ability as assessed by the Tower of London (TOL) task in homozygotic T carriers. SORL1 rs11218343 T carriage was negatively associated with processing speed as assessed by the Symbol Digit Modalities Test, *B* = −0.15, SE = 0.074, *p* < 0.05. Albeit, the effects were *not* statistically significant, Bressler et al. (2015) found a negative association between SORL1 rs11218343 T carriage and mean performance a Delayed Word Recall Test (DWRT; Immediate verbal memory), β = - 0.033, SE = 0.086, *p* = 0.703, the Digit Symbol Substitution Test (DSST; Processing speed), β = - 0.021, SE = 0.367, *p* = 0.566, and the Word Fluency Test (WFT; Executive function), β = - 0.018, SE = 0.451, *p* = 0.689. In terms of association with AD risk, Lambert et al. (2013) estimated non-zero decreased odds with regard to SORL1 rs11218343 C carriage, OR = 0.77, 95% CI [0.72, 0.82]. The finding was replicated in Brookes et al. (2018) cohort, OR = 0.78, 95% CI [0.37, 1.65].

The **ABCA7** gene is involved with the metabolism of lipids and phagocytosis (De Roeck, 2019). Carriage of ABCA7 rs3765650 risk variants may have a role in increased amyloid deposition in the brain and greater AD risk. Relative to heterozygotes and ABCA7 TT carriers, GG carriers were negatively associated with AD risk in a Taiwanese sample, with similar effects observed among Caucasian and African-Americans (Liao et al., 2014). While Liao discusses observed interactive effects regarding ABCA7 GG and APOE ε4, there are contradictions in the literature when considering individual findings, genetic database references, and assumptions about risk/protection alignment with what is rare/common in various populations with regard to propensity for various conditions or facets of cognition.

While located in close proximity to CLU (chromosome 8), the protein coding gene **Protein Tyrosine Kinase 2 Beta (PTK2B)**, a member of the focal adhesion kinase (FAK) family, is thought to exert independent signals relevant to the development of AD (Lambert et al. 2013; Bressler et al., 2017). PTK2B plays a role in long-term potentiation at the hippocampus, a process regarded as essential to progressive changes in synaptic strength, memory, and learning, broadly. Animal models of AD-relevant roles for PTK2B polymorphism rs28834970 suggest an early role in Tau pathology (Dourlen, 2017). Lambert et al. (2013) conducted a large-scale meta analysis (LOAD N = 17,008, Control N = 37,154) and showed an association between the PTK2B rs28834970 locus and increased LOAD risk among European samples, OR = 1.10, 95% CI [1.08, 1.13]. Minor C allele carriage was associated with increased Parkinson’s risk, OR 1.84, 95% CI [1.27, 2.66], *p* = 0.001, and lower expected performance with regard to the Addenbrooke’s Cognitive Examination Revised (ACE-R), a brief screening tool of dementias, however the differences were not statistically significant, M_diff_ = 2.913, SE_diff_ = 1.569, C: 75.40 ± 16.94, No C: 78.32 ± 15.44, *p* = 0.064 (Chen et al., 2018). Such differential effect of allelic carriage at PTK2B rs28834970 is further corroborated by Chan et al. (2015) with C carriage identified as a relatively detrimental risk allele in Alzheimer’s pathology. Li et al. (2016) found statistically significant differences in PTK2B rs28834970 frequencies in subjects presenting late-onset AD and controls relative to C allele carriage among APOE ε4 carriers, suggesting copathogenic effects, LOAD: 26.43%, Controls: 20.16%, OR = 1.423, 95% CI [1.041, 1.945], *p* = 0.027.

Given the complexity of understanding genetic-cognitive findings as they reflect empirical regularities regarding stable biological characteristics from which *appreciable variance* in facets of human cognitive abilities may emerge - the present work applies a machine learning feature engineering approach to provide unmeasured genetic architecture to the predictive modeling pipeline.

### Hypotheses

The first hypothesis focuses on confirming a common empirical regularity regarding age and cognitive performance. While true variance exists, it is hypothesized that participant performance on the cognitive assessment (outcome) is, on average, impeded with increasing age (years).

The second hypothesis concerns the clusters that emerge from dimensional reduction of the genetic target features. That is, clusters (separable low-dimensional subspaces) of participants that emerge from the unsupervised learning process are *expected to meaningfully stratify variance* in performance on the cognitive assessment (outcome), providing transparency into what constitutes *membership* in a given cluster and a basis for inference from targeted, emergent, genetic interaction.

Moreover, it is assumed that standardized effect sizes for each included predictor provide meaningful estimates of the *relative magnitude* of each feature in terms of its influence on the outcome. From this, the relative influence of features, including age (years), gender, education (years), *additional cognitive tasks*, as well as targeted genetic variant features (raw and derived) provide a window into the value attributable to each facet of (assumed) associated subspace in terms of the variance of the outcome cognitive assessment, at a minimum, with regard to the participant sample and in lieu of unmodeled dimensions.

### Participants

An analysis sample was formed from a participant group taking part in the Alzheimer’s Disease Neuroimaging Initiative (ADNI). Two-hundred and ninety (290) participants constitute the analysis sample, A, including a 63%/37% gender split among those whose mean age was 71 (years), ranging from 57 to 90, and whose average level of education (years) was 16, ranging from 8 to 20. Tables sourced from the database were screened, joined, and distilled to non-missing target dimensions. All data elements are de-identified and available remotely to research applicants: https://adni.loni.usc.edu/

### Quantitative Methods

To address the inherent complexities of high-dimensional genetic architectures and isolate synergistic risk patterns, an unsupervised machine learning framework is employed. Dimensionality reduction was achieved through ***Generalized Low Rank Modeling*** (GLRM), with the resulting latent features (K = 4) subsequently stratified via ***K-means clustering*** (*K* = 3) to define distinct genetic phenotypes. The prognostic utility of these data-driven sub-populations was evaluated through a ***multiple regression*** model predicting non-missing Mini-Mental State Examination (MMSE) performance. This analysis integrated derived cluster memberships alongside established clinical and demographic covariates, including age, educational attainment (PTEDUCAT), gender (PTGENDER), marital status (PTMARRY), and handedness (PTHAND). Relative feature importance was estimated utilizing partial eta squared (*η_p_*^2^) effect sizes across all included predictors. To ensure biological interpretability of the results, centroid analysis of the GLRM subspace was conducted to identify the primary genetic targets that characterize individual cluster membership.

## Results

### Generalized Low Rank Model

Four (4) generalized low rank models (GLRM) were fitted to the input dataset, A_SNP_, exclusively to the target genetic variant columns. K (rank) was varied from 1 to 6, seeking the fewest archetypes that account for the most possible A_SNP_ reconstruction ability (via minimization of the objective function). A GLRM with rank 4 was chosen and applied to achieve the predictive concatenation of 4 numeric columns to form *A*. The first archetype explains 56% of the variance in A_SNP_; Archetypes 2 through 4 explain 12%, 10%, and 7%, respectively.

| GLRM Archetypes |  |  |  |  |  |  |  |
| --- | --- | --- | --- | --- | --- | --- | --- |
|  | clu | abca7 | picalm | bin1 | sorl1 | epha1 | ptk2b |
| 1 | 1.054 | 0.051 | 0.564 | 1.355 | 0.066 | 0.223 | 0.082 |
| 2 | -0.807 | 0.347 | 1.279 | 1.379 | 0.682 | 0.201 | 0.102 |
| 3 | 0.777 | 0.042 | 0.828 | 0.341 | 0.753 | 0.259 | 0.072 |
| 4 | 0.814 | 1.038 | 0.261 | 0.886 | 0.742 | 0.420 | 0.147 |

### Kmeans Clustering

A kmeans clustering model was fitted to the GLRM genetic latent archetype dimensions. Given the *Furthest* initiation procedure, an initial center is chosen at random, followed by the next center chosen to be the furthest away in terms of Euclidian distance, estimating 3 distinct *clusters of genetic architecture*, forming *A_Clust,_* and partitioning cluster memberships into groups of 140, 83, and 67 ADNI participants.

### Generalized Additive Model

A generalized additive model (GAM) was fitted for non-linear estimation of effects, applying splines (s) to age (years), education (years), genetic archetypes 1 & 2, and a tensor product interaction surface to education and genetic archetype 2 (ti). Linear fixed effects were specified for the rest of the predictors included (see Table 2).

**Table 1.**
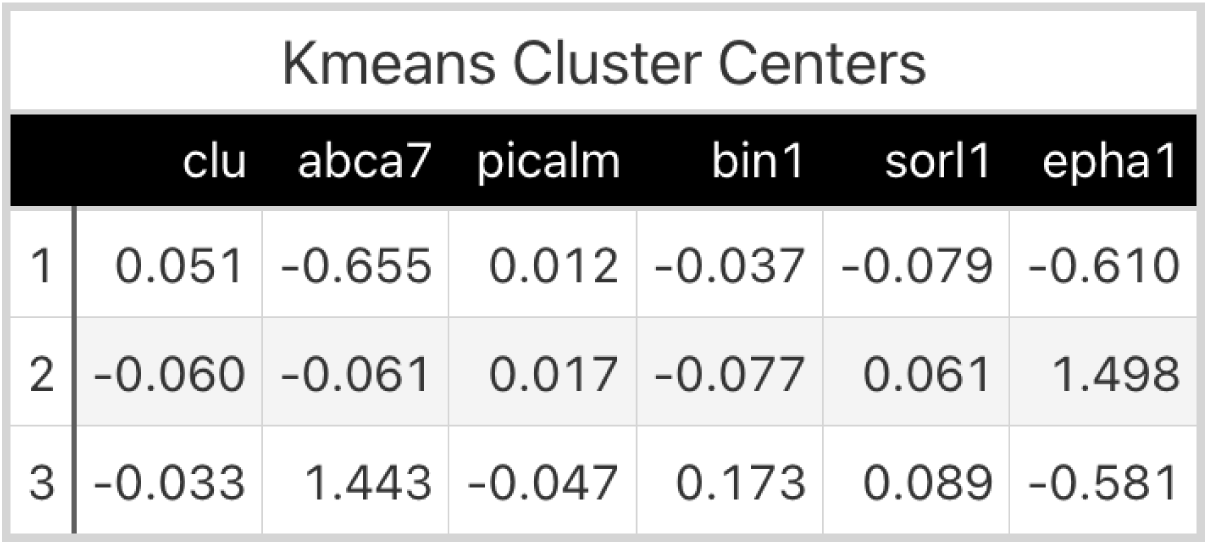

**Table 2.**

**Genetic Architecture GAM Parameter Estimates**
| *Predictors* |  |  | **MMSE** |  |  |
| --- | --- | --- | --- | --- | --- |
|  | *Estimates* | *std. Error* | *CI* | *Statistic* | *p* |
| (Intercept) | -0.24 | 0.22 | -0.67 – 0.20 | -1.06 | 0.291 |
| ABCA7 | -0.55 | 0.19 | -0.92 – -0.17 | -2.90 | **0.004** |
| EPHA1 | 0.32 | 0.35 | -0.38 – 1.01 | 0.90 | 0.368 |
| PTHAND | 0.11 | 0.16 | -0.21 – 0.43 | 0.69 | 0.491 |
| PTGENDER [1] | -0.01 | 0.12 | -0.25 – 0.22 | -0.10 | 0.919 |
| PTMARRY | 0.04 | 0.05 | -0.05 – 0.13 | 0.96 | 0.338 |
| genetic cluster [1] | -0.20 | 0.40 | -0.98 – 0.58 | -0.51 | 0.613 |
| genetic cluster [2] | 0.68 | 0.25 | 0.19 – 1.18 | 2.71 | **0.007** |
| Smooth term (age years) |  |  |  | 5.05 | **<0.001** |
| Smooth term (PTEDUCAT) |  |  |  | 4.49 | **0.008** |
| Smooth term (Arch1) |  |  |  | 0.12 | 0.730 |
| Smooth term (Arch2) |  |  |  | 1.00 | 0.410 |
| ti(PTEDUCAT,Arch2) |  |  |  | 2.72 | **0.020** |
| Observations | 290 |  |  |  |  |

Standardized partial effect sizes are displayed in Table 3, suggesting the non-linear spline specified for age stands as the highest magnitude weight to infer influence onto the cognitive assessment outcome. The non-linear effect of age on MMSE performance is estimated statistically significant, (*F* = 5.048, *p* < 0.001). Similarly, education is estimated non-zero and positive, (*F* = 4.486, *p* = 0.008). Risk variant carriage for ABCA7 is estimated non-zero and negative, (*t* = −2.896, *p* = 0.004). Interestingly, a synergistic boosting effect is observed for the tensor product interaction surface regarding education and genetic archetype 2 and displayed in Figure 2. The Figure 2 bottom-right quadrant colored plot, indicates data sparsity in dark gray as well as the MMSE boosting effect in the top-right (red), for those with the most education and highest positive score with regard to genetic archetype 2, (*F* = 2.717, *p* = 0.02). Moreover, genetic cluster membership is estimated, with *cluster* 2 estimated statistically significant relative to cluster 3 (reference), in terms of its expected influence on the cognitive assessment outcome, (*t* = 2.705, *p* = 0.007). Given the non-linear effects estimated, the reader should consider Figure 2 and the displayed heterogeneity of effects estimated instead of relying on the direction and magnitude of the statistics provided, alone. Sixteen percent (16%) of the variance (*R^2^*) in MMSE is explained by the model with a *highly* targeted genetic array and without the addition of neuroimaging or non-genetic biomarkers.

**Figure 1.**
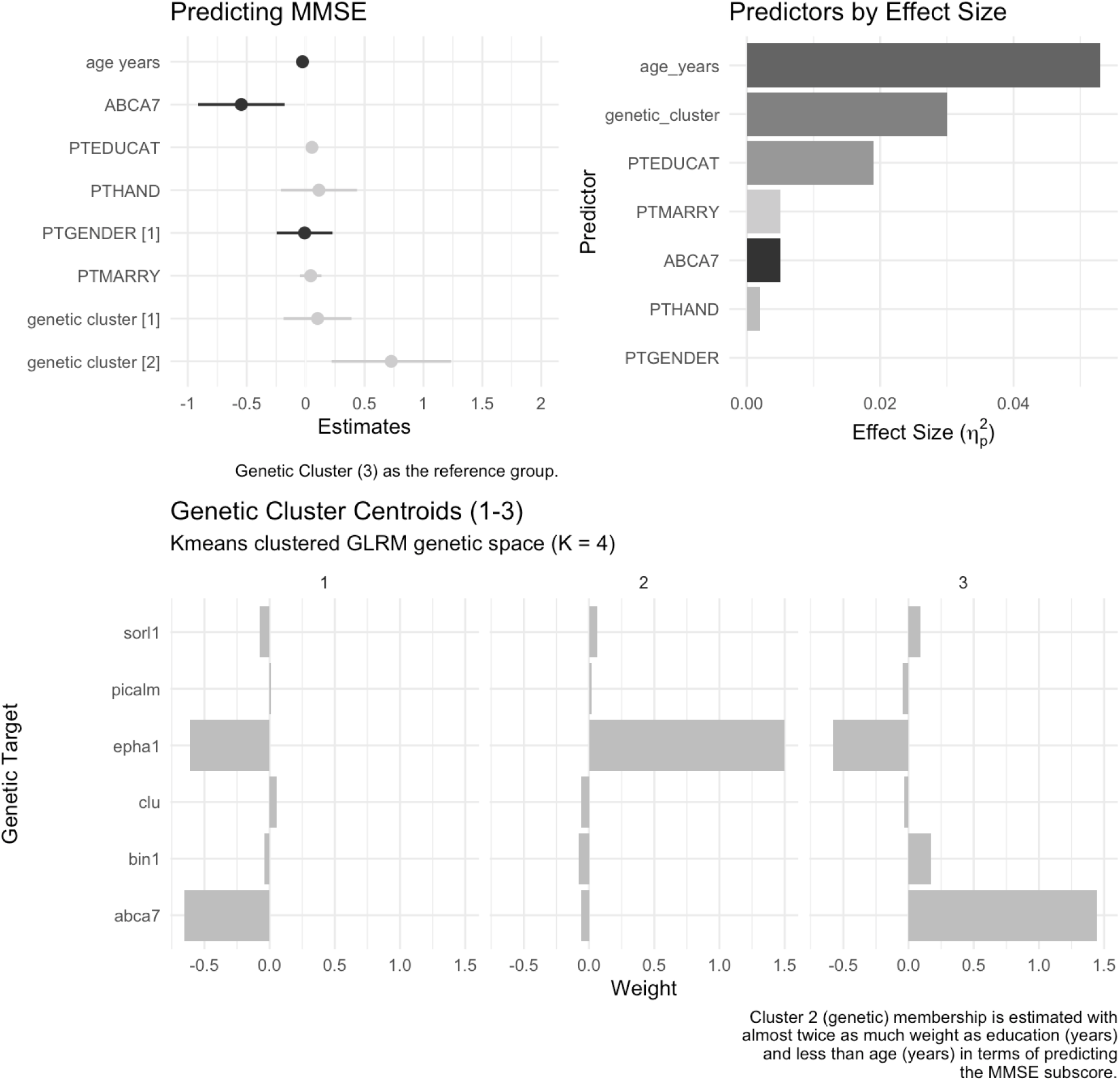

**Figure 2.**
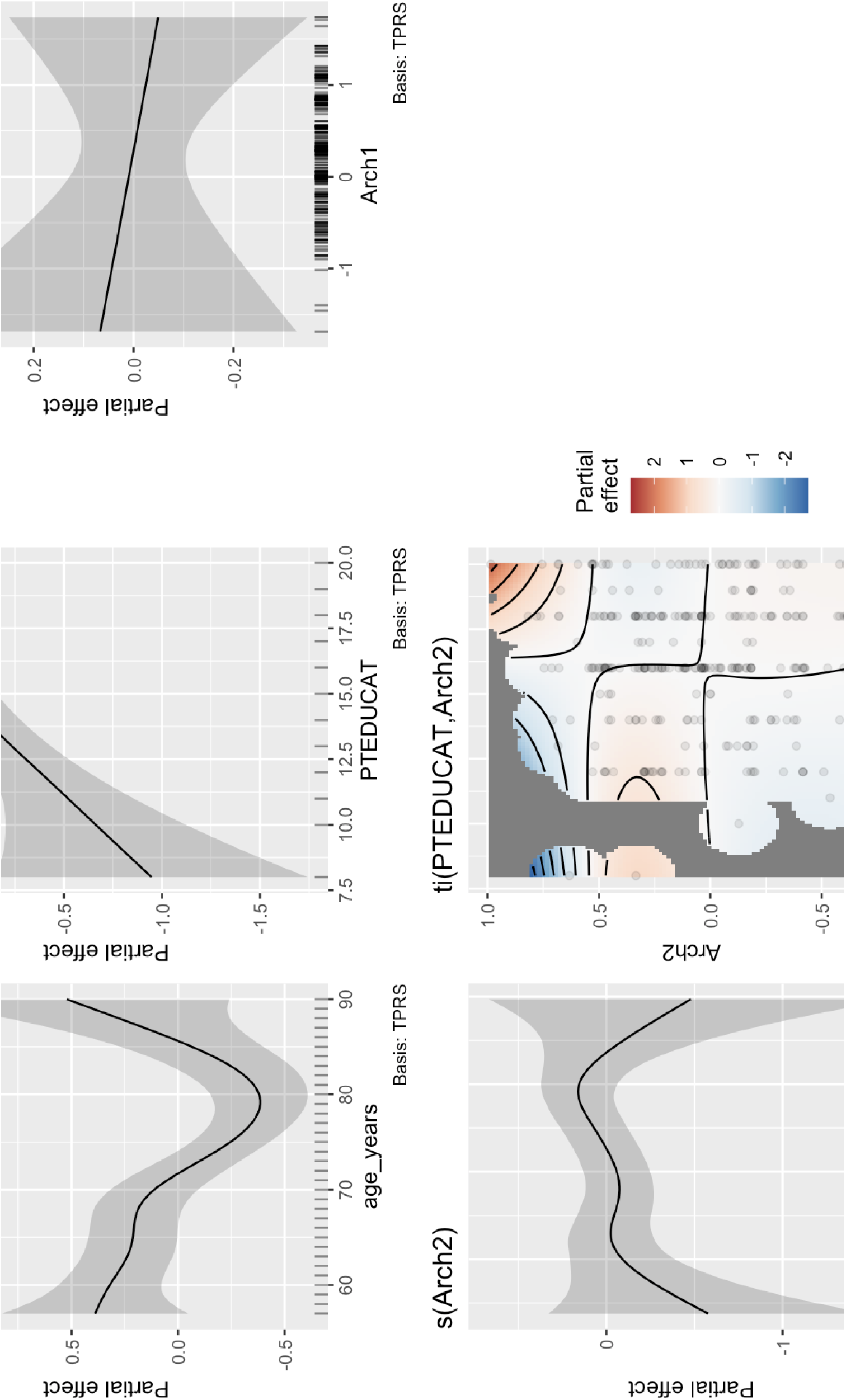

**Table 3.**

| GAM Predicting MMSE |  |  |
| --- | --- | --- |
|  | Effect Size | 95% CI |
| s(age_years) | 0.085 | [0.02106 - 0.14153] |
| ti(PTEDUCAT,Arch2) | 0.050 | [0.00107 - 0.09432] |
| s(PTEDUCAT) | 0.042 | [0.00389 - 0.09277] |
| ABCA7 | 0.031 | [0.00314 - 0.08166] |
| genetic_cluster | 0.028 | [0.00028 - 0.07404] |
| s(Arch2) | 0.018 | [0 - 0.04295] |
| PTMARRY | 0.003 | [0 - 0.03097] |
| EPHA1 | 0.003 | [0 - 0.02977] |
| PTHAND | 0.002 | [0 - 0.02551] |
| s(Arch1) | 0.000 | [0 - 0.01823] |
| PTGENDER | 0.000 | [0 - 0.00875] |
| Effect size reflect partial eta squared. |  |  |

### Limitations

Certain caveats regarding the present findings warrant further consideration. Primary among these is the demographic composition of the ADNI cohort, which remains heavily weighted toward non-Hispanic white participants with high educational attainment. Thus, the observed non-linear interactions between education and the *Arch2* architecture require replication within broader, community-derived samples to confirm generalizability. Additionally, while the MMSE serves as a standard clinical baseline, its relative lack of granularity may induce ceiling effects in high-functioning groups. Future iterations of this unsupervised GLRM pipeline might benefit from targeting longitudinal trajectories or multidimensional cognitive ability factors. Lastly, while the synergistic effects identified via the Generalized Additive Model are estimated statistically non-zero, the exact biological mechanisms by which genetic clusters interface with constructs such as cognitive reserve remain a subject for further research.

## Discussion

The present finding regarding a statistically significant main effect for ABCA7 risk variant carriage (negatively influencing variance in participants in terms of cognitive assessment performance) supports Andrews et al. (2016) finding regarding an effect of G allele carriage relative to others. Risk carriage demonstrated relative reduction in performance with regard to ABCA7 rs3765650 in immediate recall tests (short-term memory), *B* = −0.1, SE = 0.045, *p* < 0.05, however, a positive effect was shown after controlling for age with regard to the *Symbol Digit Modalities Test*, an assessment of processing speed, *B* = 4.6, SE = 1.5, *p* < 0.01. Given the present work’s inclusion of age in the GAM genetic architecture model and statistically significant, negative estimated effect on the global cognition assessment, we might consider the immediate recall test and the MMSE to tap overlapping facets of cognition more so than tasks of speeded matching, however, this also underscores the complexity of the assessment of oblique cognitive abilities, the heterogeneous demands of individual tasks & batteries, individual differences in meeting these heterogeneous demands, within-person differences, as well as the potentially *valid* and seemingly contradictory nature in facets of genetic-cognitive research that may stratify effects even further by population, sample, and other dimensions modeled and specified in other ways.

The present work did not estimate a non-zero main effect for risk carriage of EPHA1, however, it wasn’t without reason that it was considered biologically meaningful and relevant for estimation. Two-year follow-up comparisons (Wang et al., 2015) by genotype controlling for age, gender, and APOE e4 carriage showed EPHA1 AA carriers with the least volumetric decline in the left hippocampus (not statistically significant) and increases, on average, in volume of the right hippocampus relative to AD subjects with GG or AG carriage, *p* = 0.011. With regard to these changes in the right hippocampus, while 2-year follow ups showed increases in volume for AA carriers, both GG and AG carriers demonstrated decreases in volume, on average. While *seemingly* straightforward, volumetric changes may not mean what one might be quick to assume about them; *hyper*metabolism and connective inefficiency *can* involve volumetric increases and volumetric decreases could potentially be reflective of efficiency and connectivity economy that is beneficial or neutral.

### Conclusion

The current findings demonstrate the efficacy of unsupervised machine learning in the dimensional reduction and engineering of genetic architectures, yielding interpretable, *unmeasured structures* that serve as meaningful prognostic indicators for global cognitive variance. Estimation via the generalized additive model indicated a *synergistic boosting effect* attributable to the interface between years of education and the second genetic archetype extracted via the generalized low rank model. Standardized effect sizes allow us to place prioritized awareness with regard to which modeled features are expected to contribute the most to variation in the cognitive assessment.

While the relatively high positioning of age (years) in the cognitive effect size totem (in a sense serving as a sanity check) is not surprising, the elevated positioning of the effect size estimate regarding the interaction surface of education (years) and genetic archetype 2 as well as the genetic clusters (cluster 2 relative to cluster 3) - relative to the other *non-genetic* features modeled - is compelling. This demonstrates meaningful biological contributions to variance in cognitive function. By explaining variance in MMSE outcomes using only accessible demographic data and a highly targeted genetic array, this approach provides a scalable computational method that can be deployed in healthcare and clinical trial settings to support clinicians, researchers, and triage before commitment to additional assessment.

## Data Availability

All data are available at: https://ida.loni.usc.edu

## Appendix

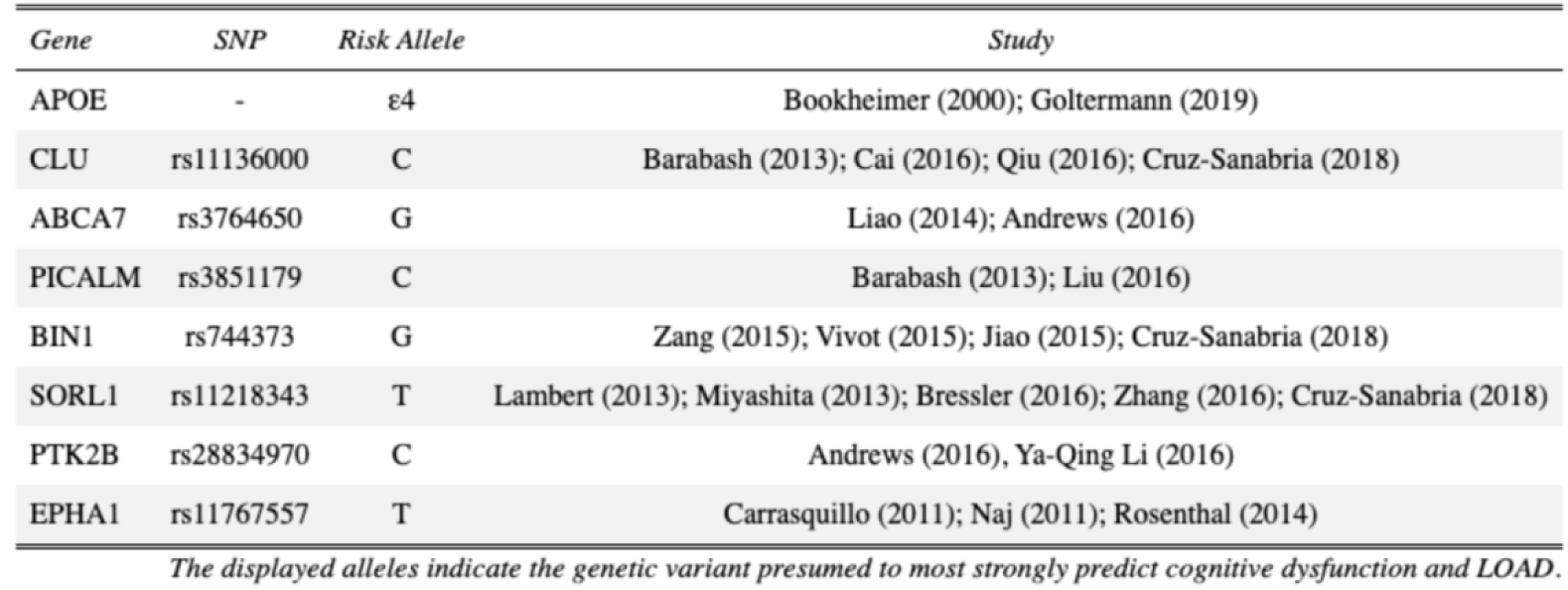

**Dementia Genes: Risk Alleles**
| *Gene* | *SNP* | *Risk Allele* | *Study* |
| --- | --- | --- | --- |
| APOE | - | ε4 | Bookheimer (2000); Goltermann (2019) |
| CLU | rs11136000 | C | Barabash (2013); Cai (2016); Qiu (2016); Cruz-Sanabria (2018) |
| ABCA7 | rs3764650 | G | Liao (2014); Andrews (2016) |
| PICALM | rs3851179 | C | Barabash (2013); Liu (2016) |
| BIN1 | rs744373 | G | Zang (2015); Vivot (2015); Jiao (2015); Cruz-Sanabria (2018) |
| SORL1 | rs11218343 | T | Lambert (2013); Miyashita (2013); Bressler (2016); Zhang (2016); Cruz-Sanabria (2018) |
| PTK2B | rs28834970 | C | Andrews (2016), Ya-Qing Li (2016) |
| EPHA1 | rs11767557 | T | Carrasquillo (2011); Naj (2011); Rosenthal (2014) |

*The displayed alleles indicate the genetic variant presumed to most strongly predict cognitive dysfunction and LOAD.*

## Notes

### Competing Interest Statement

The authors have declared no competing interest.

### Author Declarations

Source data are publicly available: https://ida.loni.usc.edu

